# Revealing patterns in major depressive disorder with machine learning and networks

**DOI:** 10.1101/2024.06.07.24308619

**Authors:** Loriz Francisco Sallum, Caroline L. Alves, Thaise G. L. de O. Toutain, Joel Augusto Moura Porto, Christiane Thielemann, Francisco A. Rodrigues

## Abstract

Major depressive disorder (MDD) is a multifaceted condition that affects millions of people worldwide and is a leading cause of disability. There is an urgent need for an automated and objective method to detect MDD due to the limitations of traditional diagnostic approaches. In this paper, we propose a methodology based on machine and deep learning to classify patients with MDD and identify altered functional connectivity patterns from EEG data. We compare several connectivity metrics and machine learning algorithms. Complex network measures are used to identify structural brain abnormalities in MDD. Using Spearman correlation for network construction and the SVM classifier, we verify that it is possible to identify MDD patients with high accuracy, exceeding literature results. The SHAP (SHAPley Additive Explanations) summary plot highlights the importance of C4-F8 connections and also reveals dysfunction in certain brain areas and hyperconnectivity in others. Despite the lower performance of the complex network measures for the classification problem, assortativity was found to be a promising biomarker. Our findings suggest that understanding and diagnosing MDD may be aided by the use of machine learning methods and complex networks.

## I. INTRODUCTION

Major depressive disorder (MDD) is a common mental condition that affects over 274 million people worldwide, according to information on depression provided by the Global Burden of Disease (GBD) study of 2019 [1]. It is ranked as the second most significant contributor to the worldwide disease burden, measured in disability-adjusted life years, spanning both developed and developing countries [2]. MDD is a disabling condition marked by the presence of at least one distinct depressive episode enduring for a minimum of 2 weeks [3]. It is characterized by a persistent and pervasive low mood, loss of interest or pleasure in most activities, and a range of other symptoms that significantly impact daily functioning [4, 5].

Previous work has shown that MDD is a multifaceted disorder involving complex neurobiological mechanisms, including the interplay of nerve growth factors, neurosecretory systems, biogenic monoamines and brain-derived neurotrophic factors [6, 7]. Despite advances in understanding its neurobiology, there is currently no established mechanism that explains all aspects of the disorder [3, 4].

The diagnosis of MDD currently relies on doctor-patient communication and scale analysis, which is prone to problems such as low sensitivity, subjective bias and inaccuracy [8, 9]. Therefore, there is an urgent need for an objective, automated method capable of predicting clinical outcomes in depression, ultimately improving the accuracy of depression detection and treatment.

Recently, a large amount of data has been generated from MDD [10, 11]. Researchers have used different techniques to study MDD, such as functional magnetic resonance imaging (fMRI) [10, 12, 13], positron emission tomography (PET) [14, 15], and electroencephalography (EEG) [11, 16–19] Recently, EEG has emerged as an important non-invasive and cost-effective tool that reveals real-time electrical activity and provides insights into neural oscillations and connectivity patterns associated with MDD. In particular, researchers have identified specific patterns and changes in brainwave activity, such as changes in frequency, amplitude and connectivity, that are indicative of MDD [19–21]

These observed changes have allowed the neuroimaging community to consider analytical methods to facilitate the classification of patients for different disorders [21–24] such as autism [25], schizophrenia [26] and Alzheimer [27]. Similarly, machine learning methods have been applied to the diagnosis of MDD through the analysis of EEG data [21–24, 28, 29], recognising subtle and intricate patterns within large datasets and identifying distinct features or biomarkers [19, 30]. The application of machine learning to EEG analysis has provided a more objective and data-driven approach to diagnosis, paving the way for personalised and effective interventions.

In a recent study [24], three machine learning algorithms were used to classify individuals with MDD: support vector machine (SVM), multilayer perceptron (MLP), and the extended K-nearest neighbours (E-KNN) model. The use of all features in conjunction with the linear SVM classifier resulted in an accuracy of 93.75%. The proposed E-KNN model showed an accuracy of 93.10%, while MLP achieved an accuracy of 92.18%. Subsequently, Movahed et al. [21] used different types of EEG features derived from MDD, including statistical, spectral, wavelet, functional connectivity and nonlinear analysis methods. The Radial Basis Function Kernel Support Vector Machine (RBFSVM) classifier, which combines all feature sets, showed the best performance of all classifiers used, resulting in an average accuracy of 99%, sensitivity of 98.4%, specificity of 99.6%, f1-score of 98.9% and a false discovery rate of 0.4%.

Deep learning methods have also been applied to MDD diagnosis [31–36]. For example, Mumtaz and Qayyum [31] presented two approaches using deep learning techniques to diagnose depression. Initially, they proposed a convolutional neural network (CNN), followed by a model that incorporated both CNN and long short-term memory (LSTM) techniques. The classification accuracies reported for the CNN and CNN-LSTM methods were 98.32% and 95.97% respectively.

Several other studies have been conducted on the classification of MDD using EEG-derived data. Table I provides a list of papers along with the respective results of each investigation, including details of the machine learning and deep learning classifiers, providing a comparative overview of their performance.

**TABLE I.** Summary of results on the performance of machine learning and deep learning methods for MDD classification using EEG time series.

| Reference | Year | Data | EEG data | Methods | AUC | Accuracy | Recall | Precision | f1-Score |
| --- | --- | --- | --- | --- | --- | --- | --- | --- | --- |
| [22] | 2017 | 33 MDD | Power of frequency bands/<br>Alpha asymmetry | LR | - | 98.33/ | 96.66/ | - | - |
|  |  | 30 TD |  | NB/ |  | 97.6/ | 96.6 |  |  |
|  |  | 19 channels |  | SVM |  | 98.4 | 97.5 |  |  |
| [36] | 2018 | 15 MDD | Right/left hemisphere<br>time series records | CNN | - | 95.49/ | 94.99/ | - | - |
|  |  | 15 TD<br>4 channels |  |  |  | 93.54 | 91.89 |  |  |
| [31] | 2019 | 33 MDD | EEG time series | CNN/ | - | 98.32/ | 98.34/ | 99.78/ | 97.65/ |
|  |  | 30 TD<br>19 channels |  | CNN+LSTM |  | 95.97 | 93.67 | 99.23 | 95.14 |
| [23] | 2020 | 34 MDD | Power of frequency bands/<br>Alpha asymmetry | SVM/ | - | 88.33/ | 89.41/ | - | - |
|  |  | 30 TD |  | LR/ |  | 85.82/ | 73.57/ |  |  |
|  |  | 19 channels |  | NB/ |  | 86.63/ | 87.42/ |  |  |
|  |  |  |  | Decision tree |  | 87.50 | 89.28 |  |  |
| [24] | 2020 | 34 MDD | Frequency bands | SVM/ | - | 93.75/ | 94.11/ | - | - |
|  |  | 30 TD |  | MLP/ |  | 92.18/ | 91.11/ |  |  |
|  |  | 19 channels |  | E-KNN |  | 93.10 | 88.23 |  |  |
| [21] | 2021 | 34 MDD<br>30 TD<br>19 channels | Combining feature sets | RBFSVM | - | 99.0 | 98.4 | - | 98.9 |

In addition to brain signal data, researchers have recently incorporated network science as an additional method for understanding the fundamental properties of functional brain networks associated with various disorders [25–27, 37–39]. Numerous studies in the literature have examined depression through the lens of brain networks [19, 40–46], providing valuable insights into the brain’s functionality in relation to this disorder. These studies use advanced methods to analyse the intricate connectivity patterns within brain networks, with the aim of distinguishing people with MDD from healthy subjects.

For example, in an EEG sleep study [40], acutely depressed patients showed significantly lower path length and no significant changes in cluster coefficient. They also showed a lower mean level of global synchronisation and a loss of small-world features. Nevertheless, the existence of numerous conflicting findings underlines the need for additional studies and further investigation. In particular, regarding path length, some authors observed longer path lengths in depressed patients [42, 43], while others reported shorter path lengths [40, 46]. Table II provides an overview of the studies, their results and the specific complex network measures used to differentiate MDD from healthy controls.

**TABLE II.** Overview of results related to network-based features. SW: Small-Worldness, FC: Functional Connectivity, PL: Path-Length, CC: Clustering Coefficient, BC: Betweeness Centrality, GE: Global Efficiency, LE: Local Efficiency, NS: Node Strength, ND: Node Degree.

| Reference | Year | Complex network features | Findings related to MDD group |
| --- | --- | --- | --- |
| [40] | 2009 | PL, CC | Decreased PL and no significant changes in CC |
| [41] | 2017 | SW, FC | Decreased SW and increased FC |
| [42] | 2018 | PL, CC, GE, NS | Increased PL and decreased CC, GE, NS |
| [46] | 2018 | PL, CC | Decreased PL and decreased CC |
| [43] | 2018 | PL, FC | Increased PL and decreased FC |
| [44] | 2019 | CC, BC | Decreased CC and decreased BC |
| [45] | 2020 | GE, LE, BC, ND, NS | Increased GE and BC, no changes in LE, ND and NS, and a more randomized structure |

Thus, previous studies have shown that the analysis of raw EEG data and the use of complex networks are promising approaches to improve the diagnosis of MDD. In this paper, unlike previous work, we integrate machine learning with network science into a single framework to study MDD data.

Our primary goal is to diagnose MDD and understand the distinctive brain topological properties associated with this condition. We investigate the influence of reconstruction methods and machine learning algorithms on the classification process to achieve an accurate diagnosis. We verify that the Spearman correlation coefficient emerges as the most effective connectivity metric for network construction. We compute complex network measures from the connectivity matrix to determine the brain differences in MDD compared to typical development (TD). Specifically, the functional MDD brain network is characterised by increased assortativity, decreased connectivity density, and a more randomised topology. Our analysis also complements previous studies by incorporating data enrichment through multiple time windows to analyse brain signals. By integrating these different methods, metrics and machine learning algorithms, we achieve more accurate results than those reported in previous studies.

## II. DATA AND DATA PRE-PROCESSING

The MDD dataset used in this paper was obtained by [22]. It consisted of two groups of participants: 34 MDD patients (17 women and 17 men, mean age = 40.3 ± 12.9) and 30 age-matched healthy controls (9 women and 21 men, mean age = 38.2 ± 15.6). Participants were recruited from the outpatient clinic of the Hospital Universiti Sains Malaysia (HUSM), Malaysia. EEG signals were recorded with the eyes closed from 19 scalp channels including frontal (Fp1, Fp2, F3, F4, F7, F8, Fz), temporal (T3, T4, T5, T6), parietal (P3, P4, Pz), occipital (O1, O2) and central (C3, C4, Cz) regions. The Cz electrode used as a reference was removed, leaving a total of 18 electrodes. For further details see [22].

The EEG dataset was pre-processed according to the methodology described by Toutain et al. [47]. The cleaning process was performed using EEGLAB from MATLAB. A bandpass filter from 0.5 to 48 Hz was applied and the data were then segmented into 1.05 second epochs. A threshold of ±70 *μV* was used, and epochs containing artefacts above the threshold (such as those associated with eye movements or muscle activity) were removed. Only eye closed EEG data from 29 MDD patients and 28 TD subjects were used in this study.

## III. METHODOLOGY

Our methodology is illustrated in Figure 1.

**FIG. 1.**
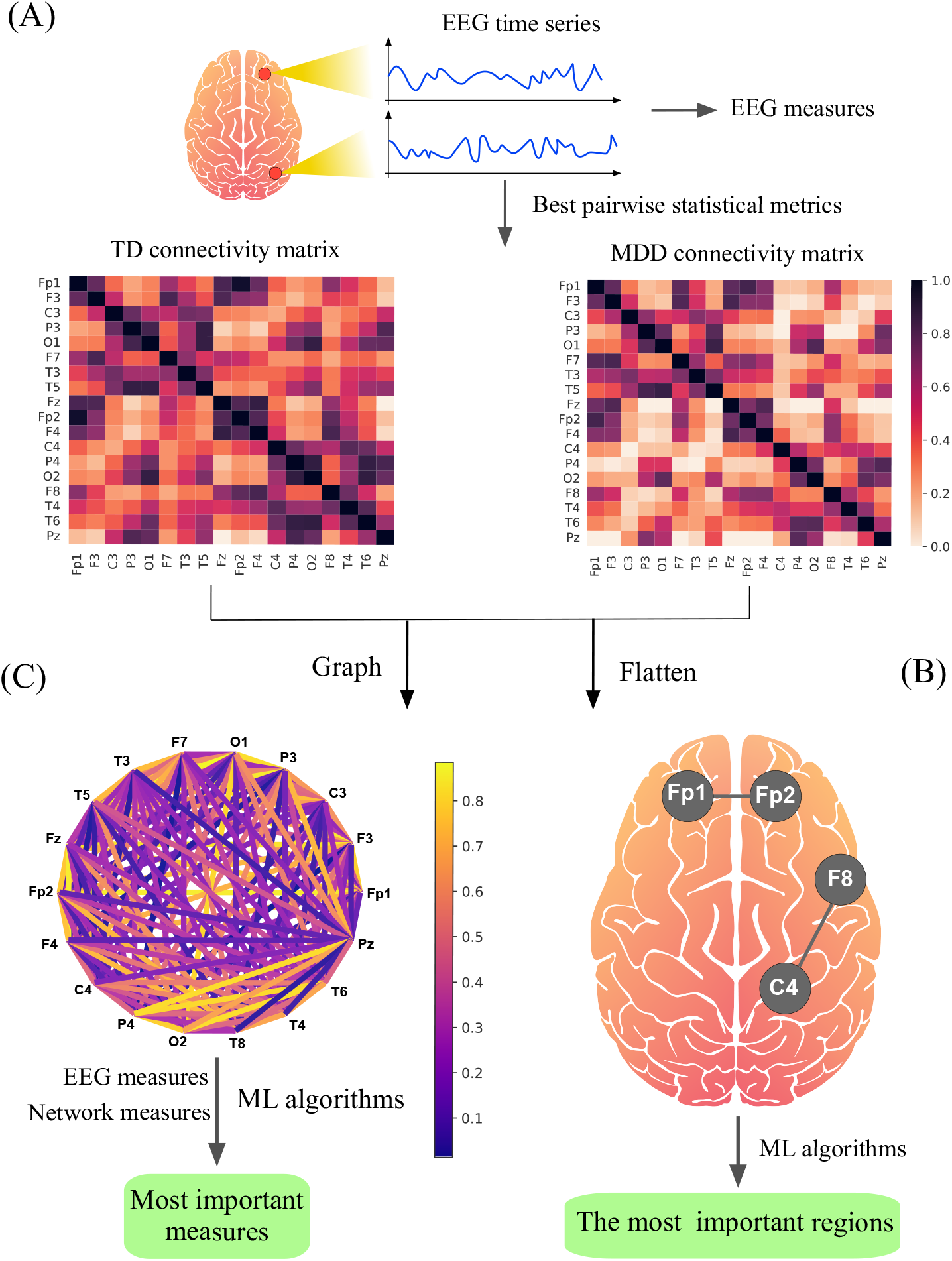
The methodology for distinguishing MDD from TD patients. In (A), EEG signals are pre-processed and the most appropriate method for constructing the connectivity matrix is identified (see subsection III A). In (B), machine learning algorithms are used to find the most significant brain regions to discriminate between MDD and TD subjects. In (C), complex network measures quantify the main topological difference between TD and MDD brains (see subsection III B).

### A. Connectivity matrices

From the 18-channel EEG time series, the connectivity matrix was constructed using Pearson Correlation (PC), Spearman Correlation (SC), Mutual Information (MI), Transfer Entropy (TE), Ledoit-Wolf Shrinkage (LW), Sparse Canonical Correlation Analysis (SCC) and Synchronization (Sync), as shown in Figure 1(A). Each matrix was then reduced to vectors and used as input to the ML algorithm, as shown in Figure 1(B). The Support Vector Machine (SVM) algorithm was used to determine the most appropriate metric for constructing the connectivity matrices.

We used SVM because it has been widely employed in the literature to classify brain disorders [48]. Its relative simplicity and flexibility make it suitable for addressing various classification problems. In addition, SVM has a lower computational cost than other methods and is known to be robust to overfitting. Furthermore, it generally shows good predictive performance even with limited sample size, which is common in neuroscience.

The dataset was divided into training and test sets, with 25% of the data constituting the test set. A k-fold cross-validation procedure was used for model selection and hyperparameter optimisation, where *k* = 10, along with the grid search method.

Once the optimal brain connectivity metric was identified, the following machine learning classifiers were used: (i) Naive Bayes (NB), (ii) Random Forest (RF), (iii) Logistic Regression (LR), (iv) Multilayer Perceptron (MLP), and (v) Extreme Gradient Boosting (XGBoosting).

Accuracy was adopted as the standard performance metric for evaluating both connectivity metrics and machine learning and deep learning classifiers. Other common metrics such as AUC, precision and recall were also included. To evaluate the performance of the classifiers, we examined the confusion matrix, the receiver operating characteristic (ROC) curve and the learning curve. Using the best connectivity metric and the best machine learning classifier, the SHAPley additive explanation (SHAP) method was used to identify the differences between MDD and TD brain regions.

### B. Complex network measures

An undirected binary graph was generated for each connectivity matrix to extract different complex network measures, as shown in Figure 1(C). The complex network measures were stored in an attribute matrix, where each column represents a complex network measure and each row represents a subject.

The following complex network measures were calculated to describe brain structure Average Shortest Path Length (APL) [49], Betweenness Centrality (BC) [50], Closeness Centrality (CC) [51], Diameter [52], Assortativity Coefficient [53, 54], hub score [55], eccentricity [56], eigenvector centrality (EC) [57], average degree of k-nearest neighbours [58] (Knn), mean degree [59], entropy of the degree distribution (ED) [60], transitivity [61, 62], second moment of the degree distribution (SMD) [63], complexity, k-core [64, 65], density [66] and efficiency [67]. These 17 complex network measures were used in previous studies [25–27, 68, 69].

In addition, the average path length within the largest community of each network was calculated. The resulting single value was added to the attribute matrix. The community detection algorithms used were fastgreedy (FC), infomap (IC), leading eigenvector (LC), label propagation (LPC), edge betweenness (EBC), spinglass (SPC) and multilevel community identification (MC). The abbreviations have been extended to include the letter ‘A’ (for average path length) to denote the respective approach (AFC, AIC, ALC, ALPC, AEBC, ASPC and AMC).

The most accurate classifier algorithm, together with the most appropriate connectivity metric, was used to classify MDD patients. Performance evaluation included confusion matrix, ROC curve and learning curve. In addition, the SHAP method was used to identify the most relevant complex network measures to distinguish the topology of the MDD brain.

## IV. RESULTS

### A. Connectivity matrices

First, we conducted a performance analysis using the entire time series. To achieve this, connectivity matrices were constructed by applying connectivity-related metrics to compute the relationships between pairwise time series. The results are shown in Figure 2.

**FIG. 2.**
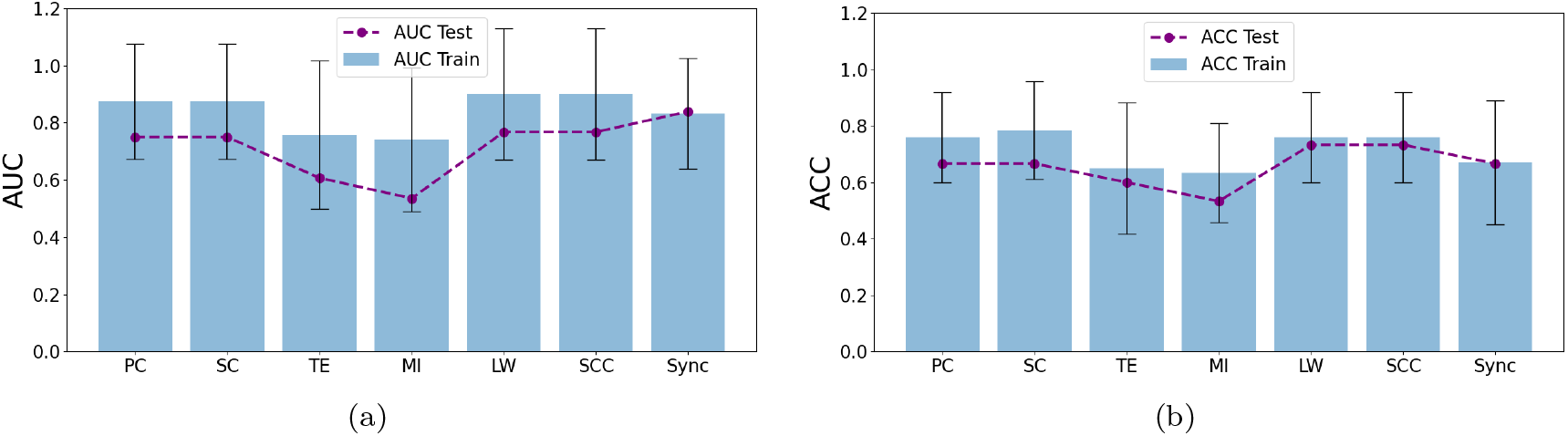
(a) AUC and (b) accuracy measures for each connectivity metric without using the sliding window.

According to Figure 2, the AUC values of the training set were higher than 0.5, which is expected for a random classification. However, the accuracy of all metrics in the training set was poor, with values below 0.79. Furthermore, there were remarkably high errors in both AUC and accuracy and poor performance of the test set for almost all metrics. These results are indicative of overfitting, suggesting that the model may have learned to fit the training data too closely and was unable to generalise well to unseen data. Increasing the size of the datasets may be necessary to address this issue.

To overcome the problem of small data sets in neuroscience, in this paper we use a sliding window data augmentation approach, similar to what was done previously in [25]. The 300-second EEG time series was divided into non-overlapping segments with window sizes of 5, 10, 20, 30, 40, 50, 60, 80, 100, 120 and 150 seconds to increase the sample data. These segments were used as input to the SVM classifier to identify the optimal window size for each connectivity metric.

To determine the optimal window size, we sought a balance between AUC and accuracy values for both the test and training sets. We also considered the error values associated with these metrics, giving preference to lower error rates.

After examining the results shown in Figure 3, it was evident that PC and SC exhibited superior performance compared to other metrics, with SC slightly outperforming PC. In particular, the most encouraging results were observed for window sizes below 60 seconds, where both training and test sets showed excellent performance with minimal variance. Based on our analysis, the optimal window size for PC and SC data was determined to be 20*s* and 10*s* respectively. With a window size of 20 seconds, we obtained 435 connectivity matrices for the MDD class and 420 for the TD class. Similarly, a window size of 10 seconds corresponded to 870 connectivity matrices for the MDD class and 840 for the TD class.

**FIG. 3.**
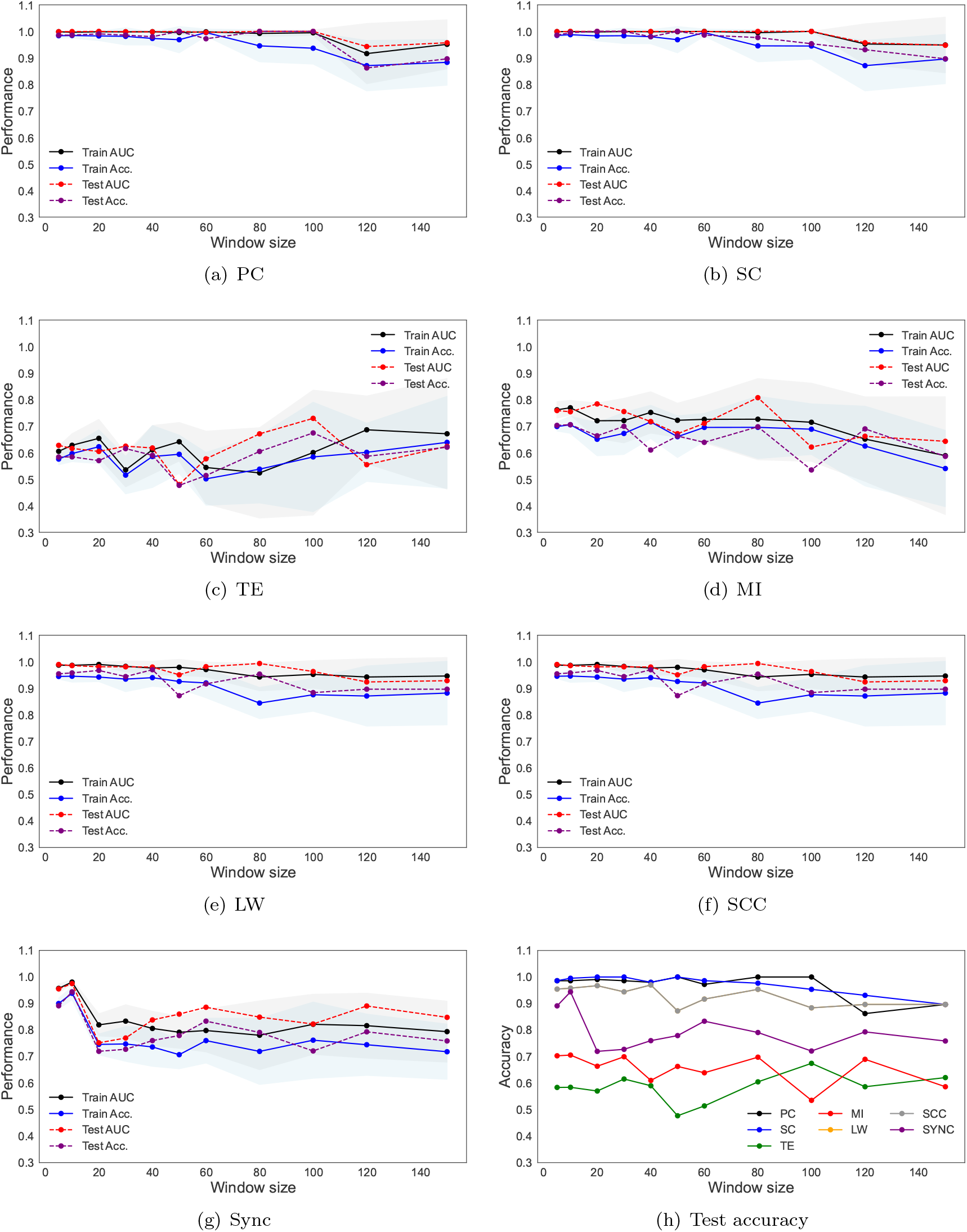
Performance measures (AUC and accuracy) for the training and test set for all the connectivity metrics with different sliding window sizes.

The other connectivity metrics displayed significant variance across almost all window sizes. The performance of TE and MI data did not yield a satisfactory result for any of the window sizes tested, and there was no discernible trend between performance and window size. Therefore, a window size of 5*s* seconds was chosen, resulting in 1740 connectivity matrices for the MDD class and 1680 for the TD class. This window size was also chosen for LW and SCC, and a window size of 10*s* seconds was chosen for synchronisation.

Table III presents the results for each connectivity matrix constructed using each of the pairwise statistical metrics, along with their respective optimal window sizes. SVM was used to determine the most effective metric for classifying MDD and TD brains.

**TABLE III.** Results obtained from SVM using different statistical methods to construct the brain network.

| Method | Window size | Subset | AUC | Accuracy | Recall | Precision |
| --- | --- | --- | --- | --- | --- | --- |
| PC | 20s | Train | $0.999 \pm 0.002$ | $0.983 \pm 0.018$ | $0.983 \pm 0.018$ | $0.984 \pm 0.016$ |
|  |  | Test | 1.000 | 0.991 | 0.991 | 0.991 |
| SC | 10s | Train | $0.999 \pm 0.001$ | $0.988 \pm 0.004$ | $0.988 \pm 0.004$ | $0.988 \pm 0.004$ |
|  |  | Test | 1.000 | 0.995 | 0.995 | 0.995 |
| TE | 5s | Train | $0.605 \pm 0.020$ | $0.576 \pm 0.022$ | $0.576 \pm 0.022$ | $0.576 \pm 0.022$ |
|  |  | Test | 0.628 | 0.584 | 0.584 | 0.584 |
| MI | 5s | Train | $0.762 \pm 0.031$ | $0.699 \pm 0.017$ | $0.699 \pm 0.017$ | $0.700 \pm 0.017$ |
|  |  | Test | 0.759 | 0.703 | 0.703 | 0.703 |
| LW | 5s | Train | $0.987 \pm 0.006$ | $0.945 \pm 0.010$ | $0.945 \pm 0.010$ | $0.945 \pm 0.010$ |
|  |  | Test | 0.990 | 0.954 | 0.954 | 0.955 |
| SCC | 5s | Train | $0.987 \pm 0.006$ | $0.945 \pm 0.009$ | $0.945 \pm 0.009$ | $0.948 \pm 0.009$ |
|  |  | Test | 0.990 | 0.954 | 0.954 | 0.955 |
| Sync | 10s | Train | $0.981 \pm 0.010$ | $0.938 \pm 0.024$ | $0.900 \pm 0.021$ | $0.900 \pm 0.021$ |
|  |  | Test | 0.975 | 0.944 | 0.891 | 0.891 |

Among the applied connectivity methods, PC (AUC = 1, 000, accuracy = 0.991) and SC (AUC = 1, 000, accuracy = 0.995) showed the best performance for the test set compared to the other metrics. However, SC was chosen over PC because of its ability to capture non-linear relationships and handle non-normally distributed data, which are common characteristics of brain connectivity data.

Three measures were used to evaluate the performance of classification models: (i) confusion matrix, (ii) ROC curve and (iii) learning curve. The results are shown in Figure 4.

**FIG. 4.**
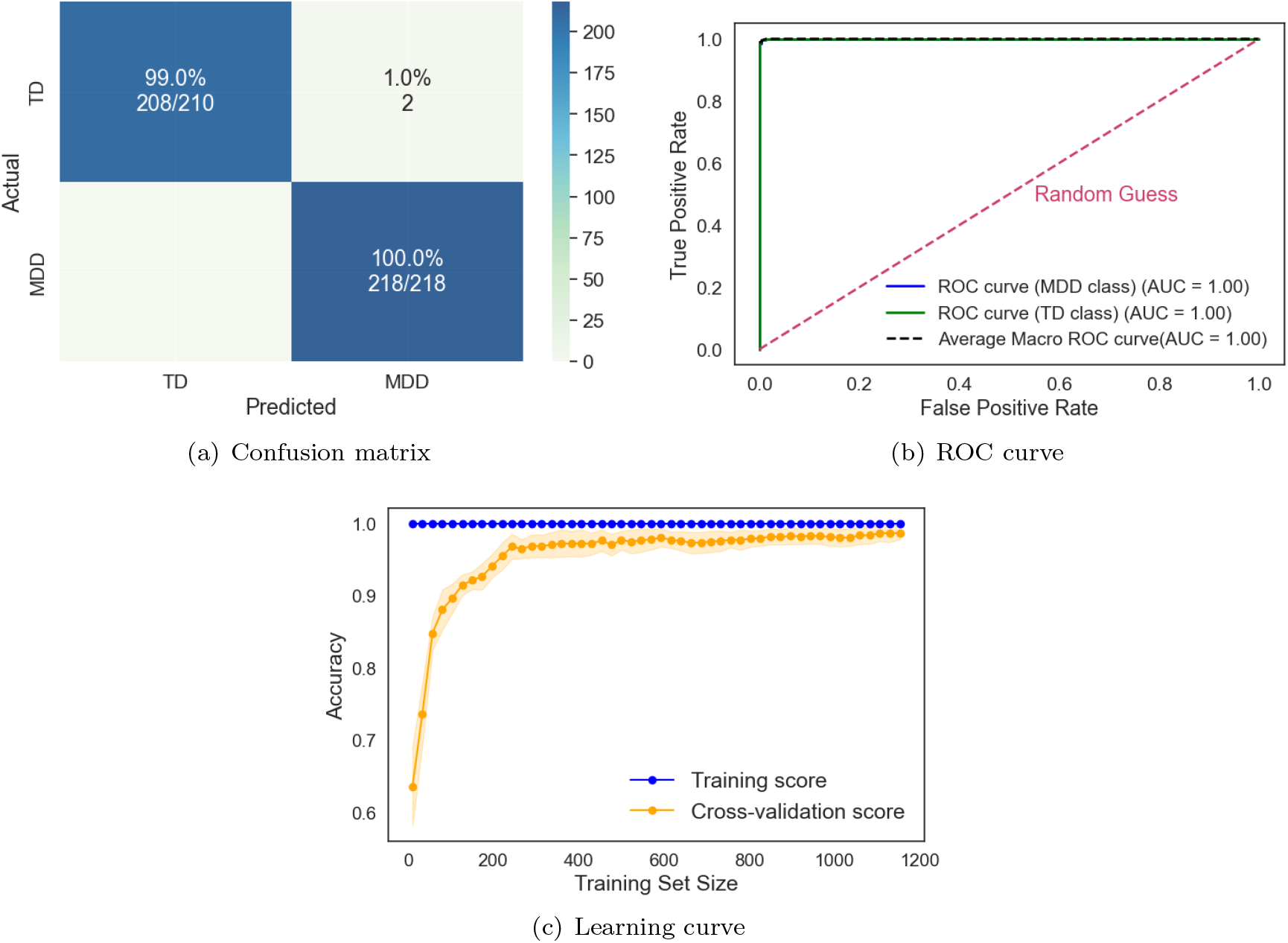
Results from the SVM classifier using as input the connectivity matrices constructed via SC and times series with a window size of 10*s*.

After identifying the best connectivity metric, we compared different machine learning methods to determine the one that provides the most accurate classification. Table IV shows the results for each machine learning method using the connectivity matrices constructed using the Spearman correlation coefficient.

**TABLE IV.**
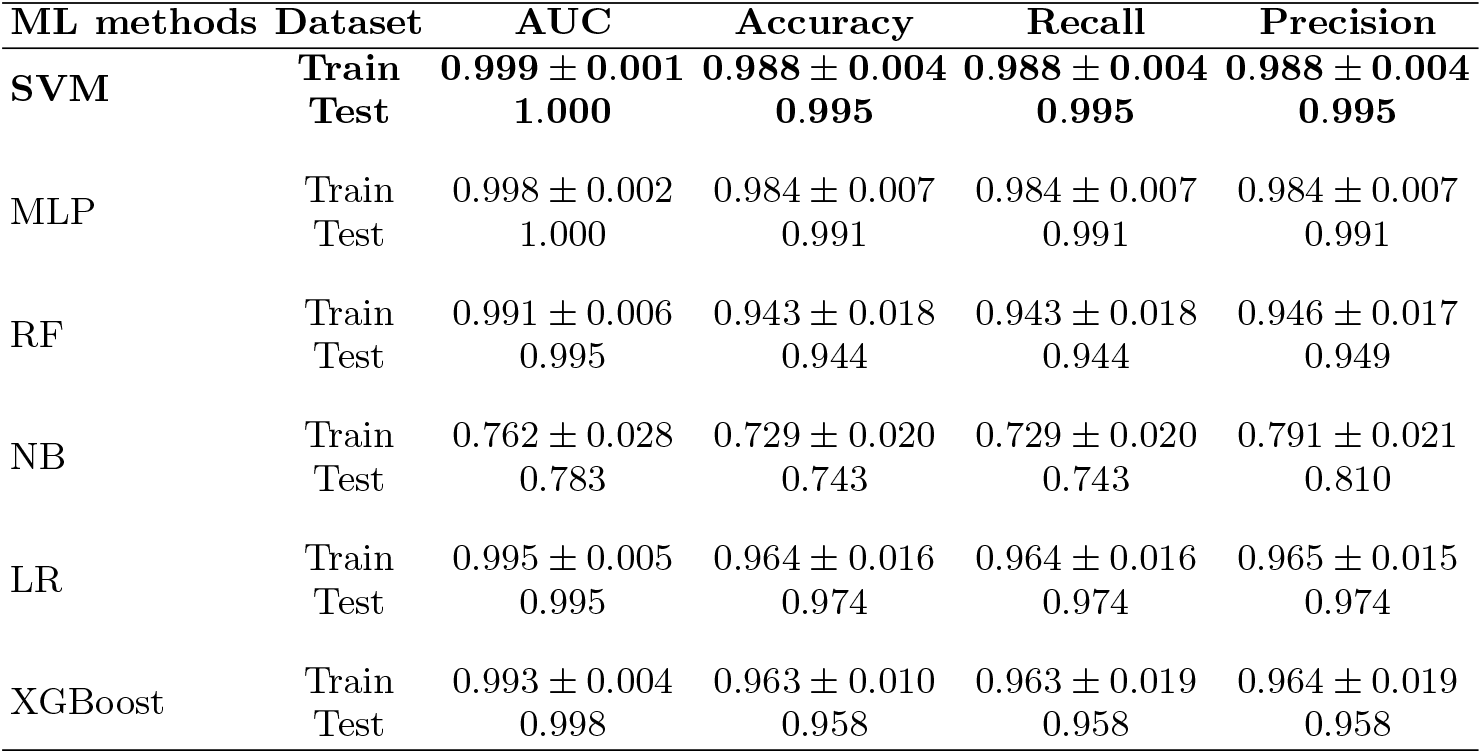
Results obtained from machine learning classifiers using the connectivity matrices constructed by employing Spearman correlation coefficient and times series with a window size of 10*s*.

| ML methods | Dataset | AUC | Accuracy | Recall | Precision |
| --- | --- | --- | --- | --- | --- |
| SVM | Train | $0.999 \pm 0.001$ | $0.988 \pm 0.004$ | $0.988 \pm 0.004$ | $0.988 \pm 0.004$ |
|  | Test | 1.000 | 0.995 | 0.995 | 0.995 |
| MLP | Train | $0.998 \pm 0.002$ | $0.984 \pm 0.007$ | $0.984 \pm 0.007$ | $0.984 \pm 0.007$ |
|  | Test | 1.000 | 0.991 | 0.991 | 0.991 |
| RF | Train | $0.991 \pm 0.006$ | $0.943 \pm 0.018$ | $0.943 \pm 0.018$ | $0.946 \pm 0.017$ |
|  | Test | 0.995 | 0.944 | 0.944 | 0.949 |
| NB | Train | $0.762 \pm 0.028$ | $0.729 \pm 0.020$ | $0.729 \pm 0.020$ | $0.791 \pm 0.021$ |
|  | Test | 0.783 | 0.743 | 0.743 | 0.810 |
| LR | Train | $0.995 \pm 0.005$ | $0.964 \pm 0.016$ | $0.964 \pm 0.016$ | $0.965 \pm 0.015$ |
|  | Test | 0.995 | 0.974 | 0.974 | 0.974 |
| XGBoost | Train | $0.993 \pm 0.004$ | $0.963 \pm 0.010$ | $0.963 \pm 0.019$ | $0.964 \pm 0.019$ |
|  | Test | 0.998 | 0.958 | 0.958 | 0.958 |

According to the table IV, all algorithms except the NB classifier showed high performance. Comparing the results for the test set, SVM gave the best performance (AUC = 1, 000, Accuracy = 0.995), closely followed by MLP (AUC = 1, 000, Accuracy = 0.991). However, SVM emerges as the preferred choice due to its ability to handle high-dimensional data efficiently, its robustness to overfitting, especially when dealing with small sample sizes, and its lower computational cost. In addition, SVM provides clear decision boundaries, which facilitates the interpretability and understanding of the classification process.

Our method achieved remarkable performance, surpassing the results found in the literature (see table I). In addition, we carefully examined potential overfitting problems and computed the AUC metric, a step not taken in other studies in the literature. The results of the confusion matrix, ROC curve and learning curve have already been shown in Figure 4.

As the performance reached almost 100%, we deliberately included random noise in the connectivity matrices of the MDD and TD groups to gain insight into the resilience of the model and its ability to recognise patterns even in the presence of perturbations, thus contributing to a more comprehensive understanding of its learning dynamics and performance under different conditions.

The noise was randomly generated from a Gaussian distribution with a mean of zero and a standard deviation ranging from 0.1 to 10. The response of the model’s performance under these conditions was measured by the average AUC and average accuracy of the test set, and the results are shown in Figure 5.

**FIG. 5.**
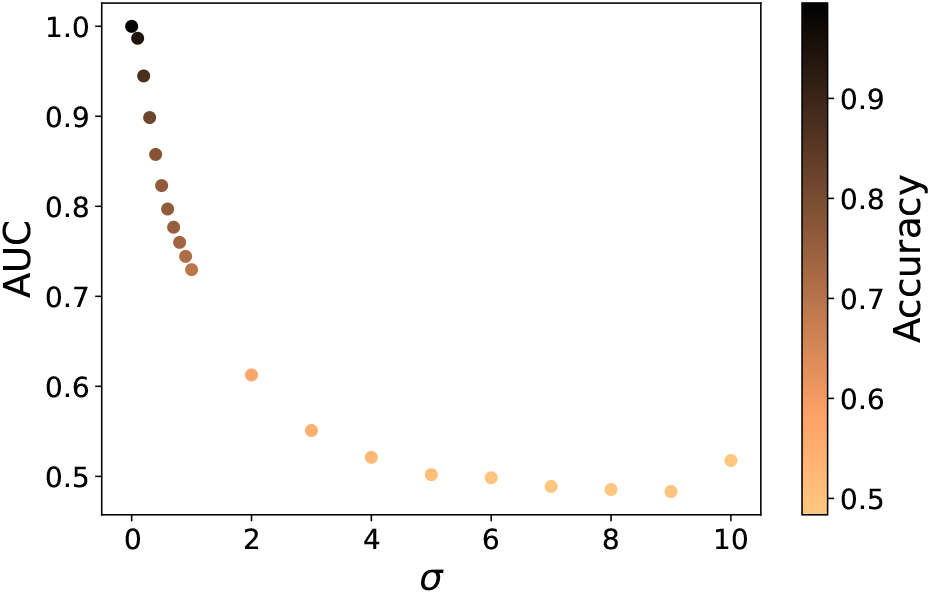
Performance of the test set for the SVM model with random Gaussian noise with a mean equal to zero and a standard deviation that varies from 0.1 to 10.

Figure 5 shows that both AUC and accuracy values decrease as the error amplitude increases, roughly following a decreasing logarithmic function. This analogous trend was also observed in the autism study by Alves et al. [25], and suggests that the model can identify patterns even in the presence of noise.

SHAP scores were then calculated to assess the importance of brain connections. Prior to this, we performed a Recursive Feature Elimination (RFE) analysis with Cross-Validation (CV) to identify the most relevant features that were essential for maintaining optimal performance with less computational cost to obtain the SHAP values. The RFECV was performed using an SVM model. From Figure 6 it can be seen that the model achieved higher accuracy with 277 features. Therefore, the full feature set was not necessary for optimal efficiency.

**FIG. 6.**
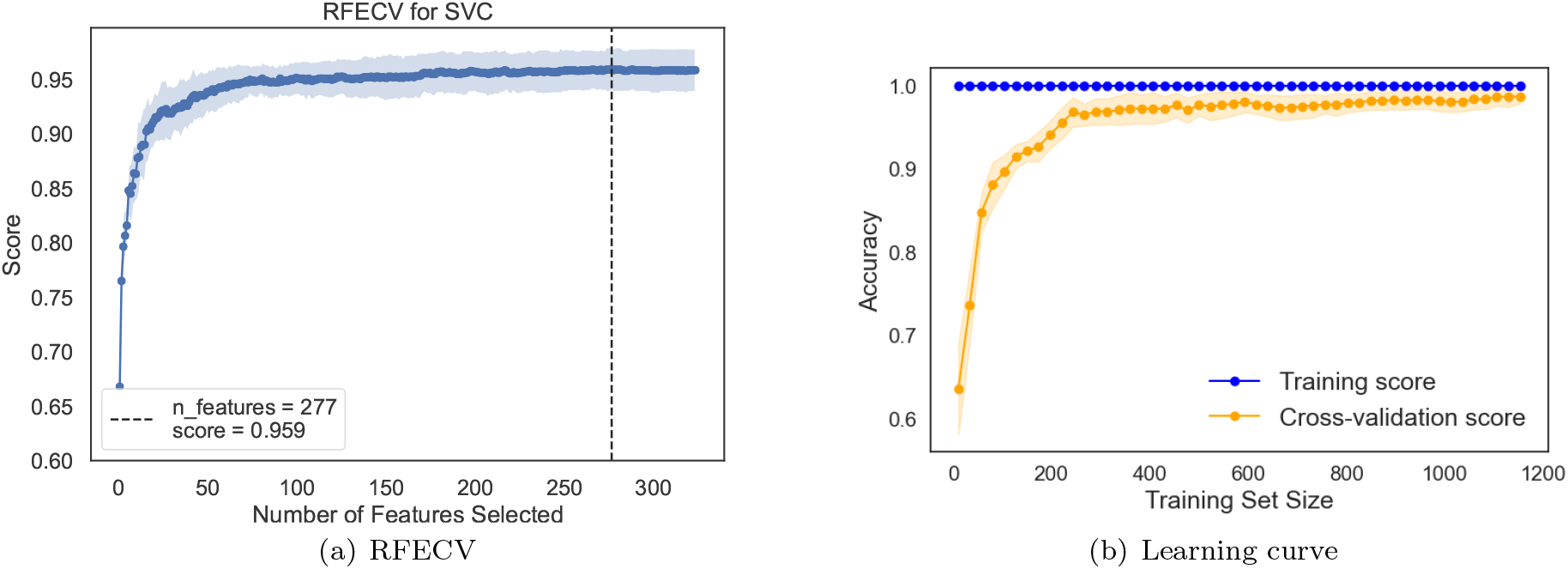
RFECV (a) and the learning curve (b) of the SVM model. In (a), it can be seen that optimal performance is achieved with 277 features. In (b), the learning curves considering the training data set (1282 connectivity matrices) are shown for the training accuracy (blue) and the validation accuracy (orange), and it can be seen that the maximum performance was obtained using the entire training set.

As shown in Figure 6(b), maximum performance is achieved using the full data set. In addition, the gap between the validation curve and the training curve remained consistent with the results obtained using all features (see Figure 4(c)), as did the variability over the validation curve. As shown in Table V, the performance remained sufficiently high. Based on these results, we used the 277 features derived from the RFECV analysis to obtain SHAP values, as shown in Figure 7.

**TABLE V.** Results obtained from SVM classifier using the 277 features selected by the RFECV.

| ML method | Subset | AUC | Accuracy | Recall | Precision |
| --- | --- | --- | --- | --- | --- |
| SVM | Train | $0.999 \pm 0.001$ | $0.988 \pm 0.004$ | $0.988 \pm 0.004$ | $0.988 \pm 0.004$ |
|  | Test | 1.000 | 0.995 | 0.995 | 0.995 |

**FIG. 7.**
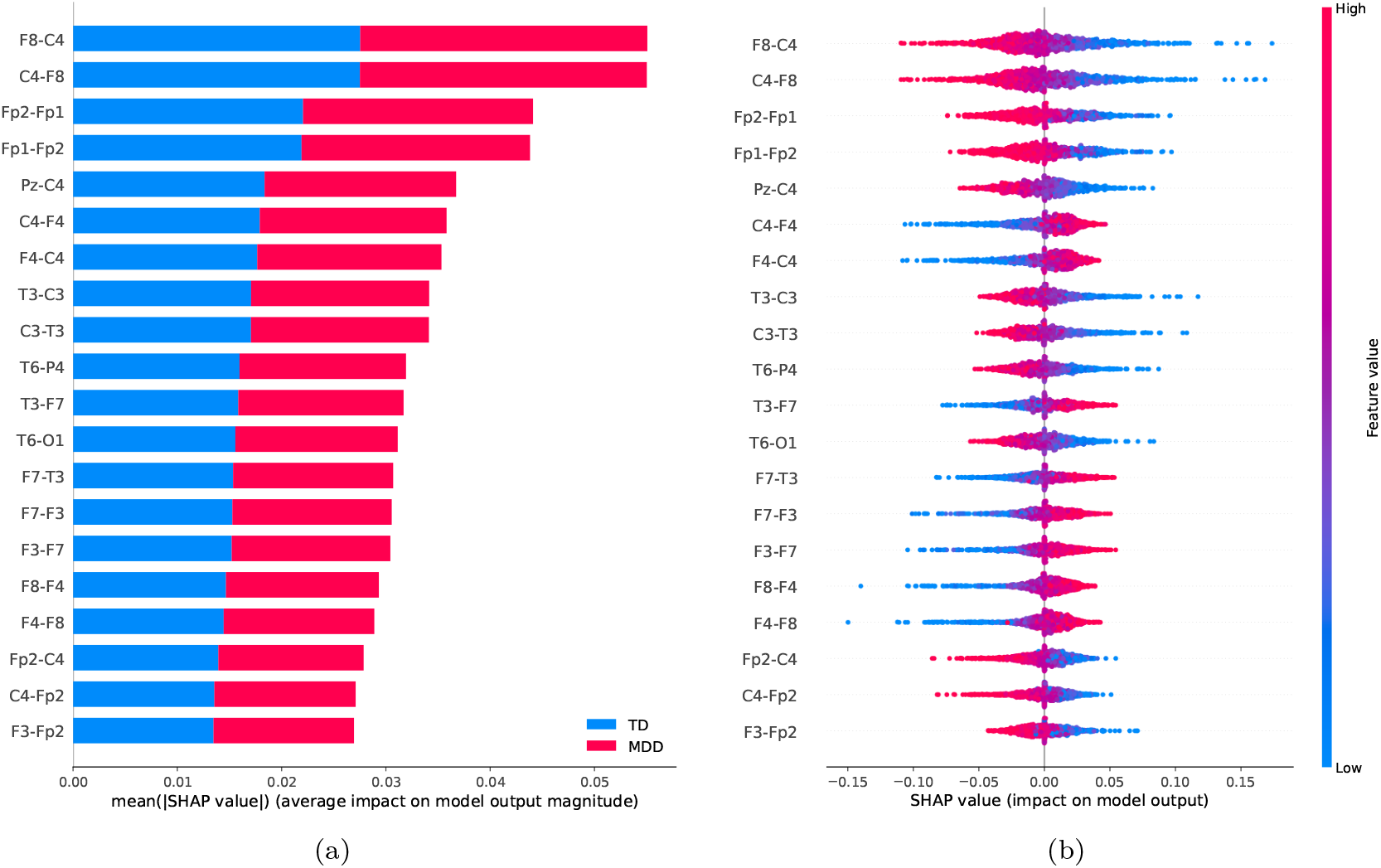
Feature importance ranking for the SVM classifier, with the brain regions ranked in descending order of importance for MDD. Figure 7(a) illustrates the average impact of each feature for the TD class and MDD class. Figure 7(b) shows the effect of each feature for the classification related to MDD class.

The most important brain connections that are crucial for distinguishing MDD patients from TD patients are ranked in Figure 7. The connection between region C4 and F8 emerged as the most important for distinguishing MDD patients from TD patients. Low PC values (blue dots) for this connection are indicative of the MDD group, whereas high PC values (red dots) are associated with the TD group. The second most important connection was between region Fp1 and Fp2. Low PC values (blue dots) for this connection characterise MDD individuals, while high correlation values (red dots) are essential for identifying TD patients.

### B. Complex network measures

We constructed the corresponding undirected binary graphs from the connectivity matrices derived using the Spearman correlation coefficient on segmented time series with a window size of 10 seconds. We then extracted various complex network measures from all graphs and stored them in a matrix. This matrix served as input to the SVM classifier, as this ML algorithm showed the best performance, as reported in the previous subsection. The results for the SVM algorithm using the complex network measures are shown in table VI.

**TABLE VI.**
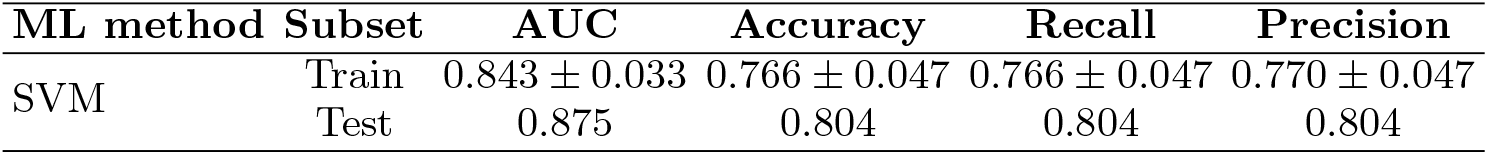
Results obtained by the SVM classifier using the complex network measures computed using the connectivity matrices constructed by the Spearman correlation coefficient with a window size of 10s.

| ML method | Subset | AUC | Accuracy | Recall | Precision |
| --- | --- | --- | --- | --- | --- |
| SVM | Train | $0.843 \pm 0.033$ | $0.766 \pm 0.047$ | $0.766 \pm 0.047$ | $0.770 \pm 0.047$ |
|  | Test | 0.875 | 0.804 | 0.804 | 0.804 |

According to the results in Table VI, the AUC and accuracy values on the test set were 0.875 and 0.804, respectively. Using network metrics for the classification problem did not lead to superior performance compared to using connectivity matrices (see Table IV). However, it did lead to an acceptable performance. The confusion matrix, ROC curve and learning curve were used to evaluate the performance and the results are shown in Figure 8.

**FIG. 8.**
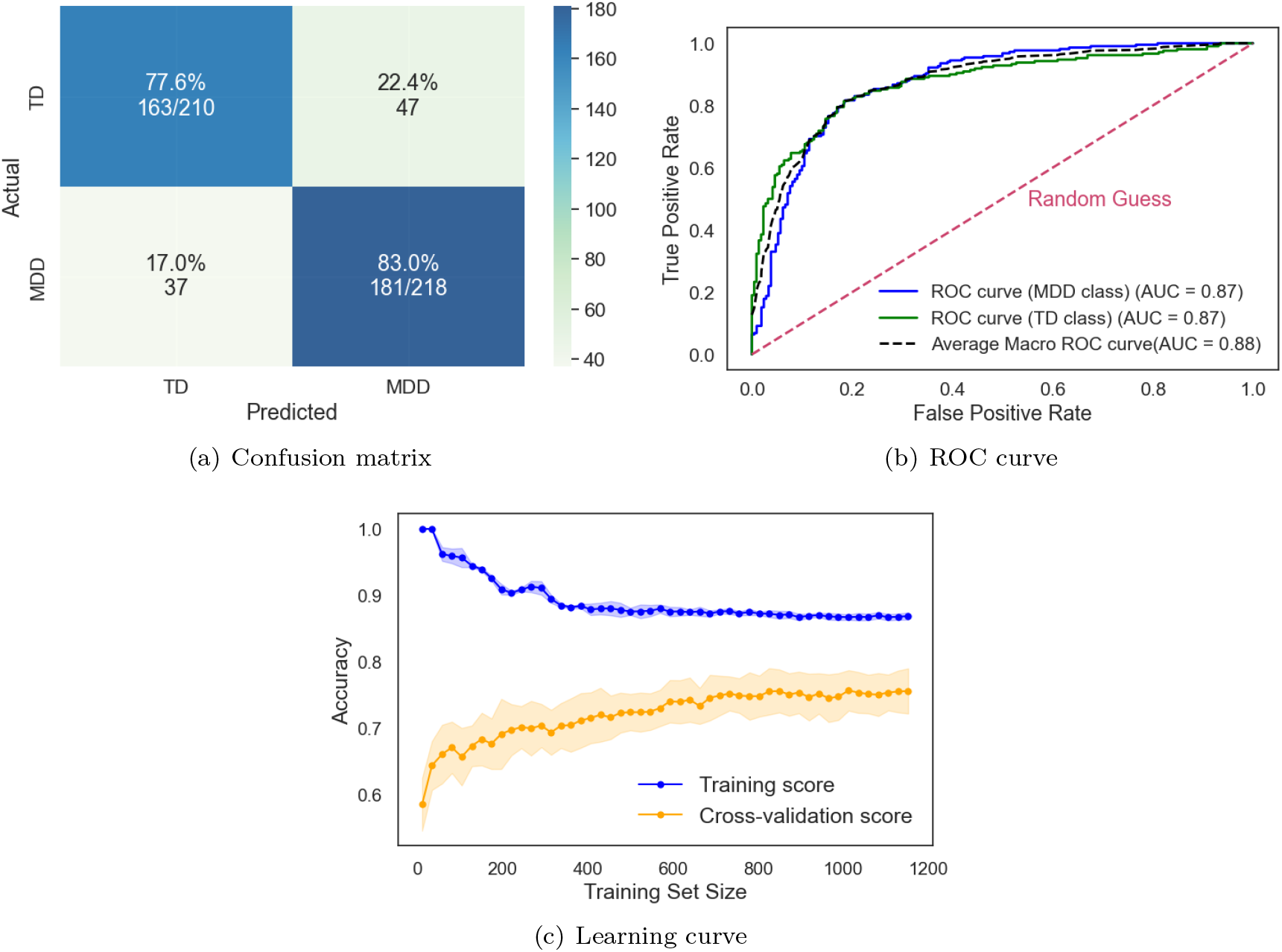
Results obtained from SVM classifier using the complex network measures that were calculated by connectivity matrices constructed through Spearman correlation coefficient with sliding window of 10s.

The influence of the complex network measures on the model’s performance was then assessed using the SHAP summary plot.

The complex network measures crucial to discriminating MDD patients from TD patients are ranked according to their global importance in Figure 9. All features are shown in order of global importance, with the first being the most important and the last being the least important.

**FIG. 9.**
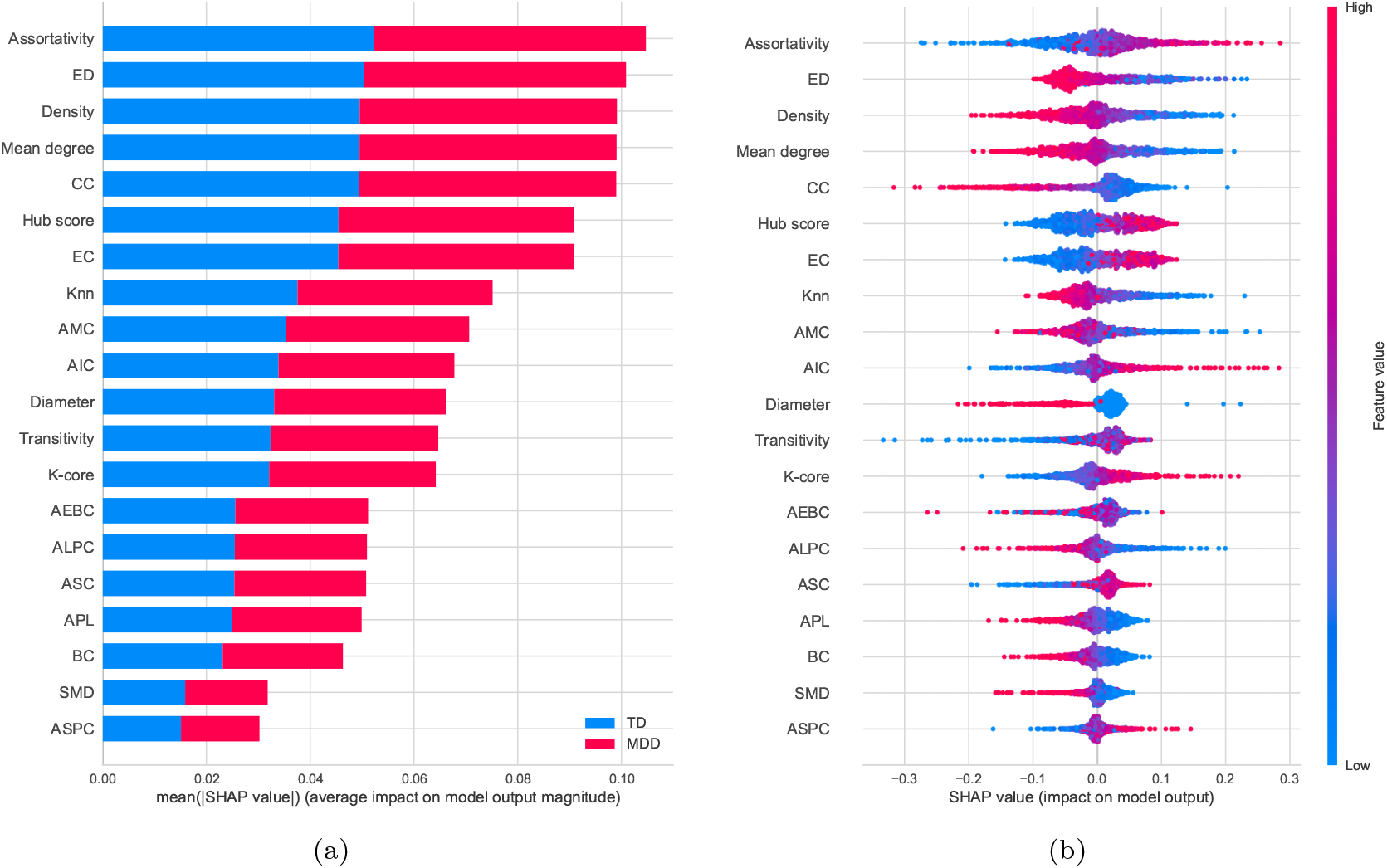
Feature importance ranking for the SVM classifier with the complex network measures ranked in descending order of importance for Major Depressive Disorder (MDD). Figure 9(a) shows the average influence of each metric for the TD class and the MDD class. Figure 9(b) shows the effect of each metric for classification in relation to the MDD class.

According to Figure 9, the most critical network measures for discriminating between MDD and TD subjects were assortativity, entropy of the degree distribution (ED) and density, in that order. High values (red dots) of assortativity have a high positive contribution to the detection of MDD patients. In contrast, low values of these variables (blue dots) have a high negative contribution to the detection of MDD patients. Low values (blue dots) of ED and Density have a high positive contribution to the detection of MDD patients. In addition, high values (red dots) of these variables have a high negative contribution to the detection of MDD patients. The same interpretation can be applied to the other variables.

## V. DISCUSSION

### A. Connectivity matrices

Among all applied connectivity methods, SC exhibited the best performance for the test set, with AUC and accuracy values of 1.000 and 0.995, respectively (see Table III). To the best of our knowledge, these results exceed those reported in the literature (see Table I). The SC metric is a nonparametric measure of rank correlation [70], widely employed as a tool in neuroscience and brain research, with consistently favourable results [71–74]. It plays a crucial role in the study of brain disorders by assessing the monotonic relationship [75] between brain regions, that is, as the signal of one region increases, so does the signal of the other region, or as the signal of one region increases, the signal of the other region decreases.

In addition, SC assesses the strength and direction of a monotonic association, allowing complex, non-linear dependencies between brain regions to be captured [76]. This suggests that the relationship between brain activities associated with depression may tend to become monotonic, not necessarily linear.

The confusion matrix (Fig. 4(a)) clearly shows that the SVM model achieves a 99% of probability in distinguishing the TD class from the MDD class and also indicates a 100% of probability that the SVM model can distinguish the MDD class from the TD class. This indicates that sensitivity (true positives) slightly exceeds specificity (true negatives), suggesting that the SVM model using connectivity matrices as input is marginally better at identifying patients with MDD than those with TD. The area under the ROC curve (Fig. 4(b)) is equal to 1.00 for both classes, indicating that the SVM model can discriminate perfectly between the TD and MDD classes.

The learning curve analysis presented in Figure 4(c) reveals some key insights. The training score remains high regardless of the size of the training set. Conversely, the test score increases with the size of the training data set, indicating that performance improves with more data until it reaches a plateau. This indicates that it is not useful to acquire new data, as the generalisation performance of the model will not increase further. There is also a relatively small gap between the training and cross-validation scores when considering the entire dataset, indicating that the model generalises well and has a good bias-variance trade-off.

Alternative machine learning methods were explored to assess whether there was an improvement in performance. As shown in table IV, both SVM and MLP gave satisfactory results. However, SVM produced slightly better results. Considering this analysis, SVM was selected as the ML method for the subsequent steps.

Although the classification performance achieved high values, the results from the insertion of random noise (Figure 5) suggest that the model can identify patterns even in the presence of noise. In other words, the model generalises to noisy data and learns the underlying patterns.

Based on the SHAP summary plot (Figure 7), the critical discriminator between MDD and TD patients was the connection between region C4 and F8, which was ranked as the most significant. This connection was followed by the connection between regions Fp1-Fp2. The SHAP plot showed that the most critical connections were located in both the right and left hemispheres of the brain, particularly in the frontal lobe. These results were consistent with previous findings in the literature, such as in [16, 77, 78].

In particular, low SC values in C4-F8 and Fp1-Fp2 connections contributed to the identification of the MDD group. The schematic brain plot in Figure 10 illustrates the key connections associated with altered connectivity in the neural network structure of individuals with MDD. The C4 electrode is positioned in a brain region that is associated with the motor cortex and voluntary movement control [81, 82], which is involved in the planning, control, and execution of voluntary movements [83]. The F8 electrode is positioned in the right frontal area of the brain. This region involves various cognitive functions, such as attention, decision-making, social skills, emotional processing, and social cognition [84–87]. Results from the literature suggest that the frontal lobe plays a vital role in depression [77, 86].

**FIG. 10.**
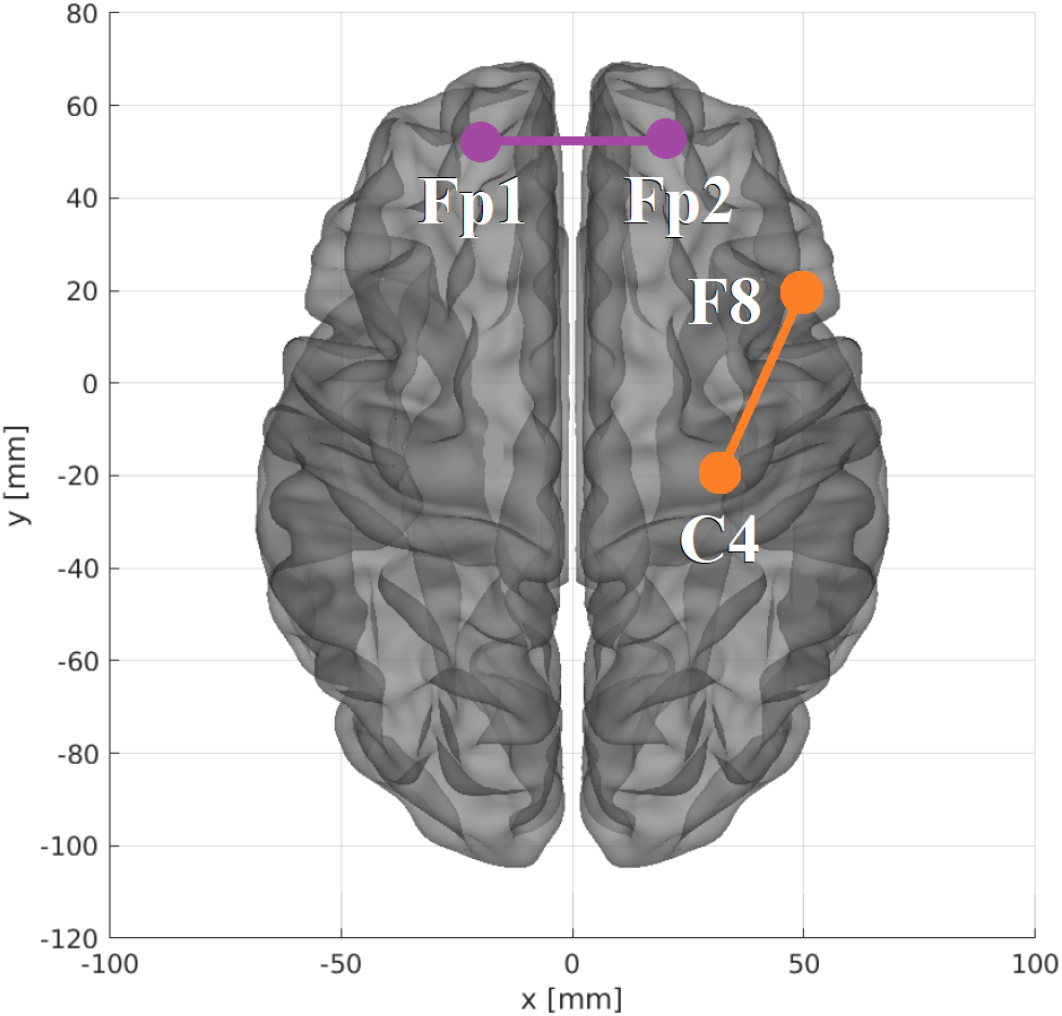
The most relevant connections are found in the two-dimensional schematic (ventral axis), highlighted in descending order of importance in orange and purple. The brain plot was developed using the Braph tool [79], based on the coordinates in [80].

Disruption in C4-F8 brain connectivity in individuals with depression may suggest deficits in attention, working memory, decision-making, and problem-solving. This observation is in accordance with the findings of the literature [88, 89], where the frontoparietal was shown to exhibit lower functional connectivity in patients with MDD than in healthy participants.

Frontal lobe dysfunction has been reported in patients with MDD [72, 86, 90]. In [72], the authors identified that, in addition to the frontal lobe, the central lobes are also involved in the impairment associated with MDD. This suggests that the fronto-central regions could serve as reliable indicators of attention deficits in MDD patients, which appears to be linked to the severity of subjective depressive symptoms. Our results are consistent with these previous findings in the literature. Additionally, a recent study on mental disorders with suicide [91] found impairment in the fronto-central regions of the suicidal group, suggesting that the brain’s electrical activity in these regions may be damaged, leading to an increased risk of suicide in mental disorders.

The Fp1 electrode is located over the left frontopolar lobe and is involved in cognition, working memory and perception [92]. The Fp2 electrode is placed over the right frontopolar cortical area and is related to affective processing and social cognition [92]. A disruption in connectivity between FP1 and FP2 could lead to cognitive dysfunction, including difficulties with concentration, memory and decision-making, which are often seen in people with MDD. It may also contribute to difficulties in regulating emotions, leading to symptoms such as persistent sadness, irritability and mood swings, which are common in MDD. These findings are consistent with the literature [93].

Figure 7 illustrates the impact of MDD on both hemispheres of the brain, in line with findings in the literature [22, 77, 94–9 In addition, Mohan et al. [97] highlighted the central brain region, particularly C3 and C4, as highly effective in detecting depression. Furthermore, Yang et al. [98] highlighted the importance of analysing the MDD brain by considering a combination of frontal, temporal and central lobe regions, confirming some of the critical regions identified in our study.

### B. Complex network measures

As can be seen from the results presented in Table VI, using network metrics for the classification problem did not yield comparable or better performance than using connectivity matrices. The AUC and accuracy results for the test set were 0.875 and 0.804, respectively (see table IV).

The confusion matrix (Fig. 8(a)) shows that the SVM model achieved an accuracy of 77.6% in discriminating the TD class from the MDD class and 83.0% in the differentiation of the MDD class from the TD class. This means that the sensitivity exceeds the specificity, suggesting that the SVM model, when using the attribute matrix as input, is also better at identifying patients with MDD than those with TD. The ROC curve (Fig. 8(b)) shows an 87% of probability that the SVM model can discriminate between the TD and MDD classes.

Regarding the learning curve (Fig. 8(c)), we can see that the training score is very high when using few samples for training, and it decreases as the number of samples increases until a certain amount of data, after which it stabilizes. In contrast, the test score is initially low and increases with the addition of more samples. Besides, there is a significant difference between training and validation accuracy.

The results indicate that complex network measures did not provide accurate results compared to connectivity matrices. This finding suggests that the differences between the brains of healthy people and those with MDD are very subtle and not captured by the measures we used. Subtle differences between the brains of controls and people with mental disorders have been observed before [99].

Despite that, the investigation regarding complex network measures offers valuable insight to uncover significant characteristics of the topology of the MDD brain and how it is discerned from the TD brain. These measures provide an understanding of the intricate connectivity patterns within the brain network, allowing for the identification of key features that contribute to the manifestation of MDD. Hence, we examined the impact of complex network measures on the model’s performance using the SHAP summary plot.

As shown in the SHAP summary plot (Figure 9), assortativity emerges as the primary discriminator between MDD and TD patients, ranking as the most significant complex network measure. ED and density are also relevant features for the diagnosis of MDD. The assortativity coefficient is a topological measure that refers to the tendency of nodes to connect to others with similar characteristics [54, 100]. Specifically, it measures the correlation between the degrees of neighbouring nodes in the network. From the SHAP summary plot, high values of assortativity are associated with the MDD group. This result contrasts with the literature, which found no difference in assortativity values between MDD and TD [101] or lower assortativity in MDD patients [102]. Higher assortativity tends to show a higher degree of modularity and leads to more robust networks, which has already been reported in the literature [45, 46].

The entropy of the degree distribution (ED) is an average measure of the heterogeneity of the network and can be used to quantify network complexity [103]. A high entropy means that there is a wide range of node degrees in the network, indicating a more heterogeneous structure. Our results suggest that MDD brains present smaller values of ED, indicating a less structured topology. Our results are also consistent with [44–46, 104].

We also observed a lower density value in MDD brains, as indicated by the SHAP summary plot. Density quantifies the extent to which nodes in the network are connected to each other [105]. A density of 0 indicates a completely disconnected network, while a density of 1 represents a completely connected network. Networks with higher densities often have more robust connectivity, while networks with lower densities may be more sparse, with fewer connections as seen in functional MDD brain network.

## VI. CONCLUSIONS AND FUTURE WORK

Our research helps to understand the characteristics of MDD brains. The analysis of different approaches to construct the connectivity matrix showed that the Spearman correlation coefficient, especially with a sliding window of 10 seconds, had the highest accuracy in discriminating between MDD and TD subjects. The SVM model was the best classifier. Our results showed an AUC of 1.00 and an accuracy of 0.993 for the test set. This performance exceeds the results documented in the literature.

From the SHAP summary plot, the right fronto-central connection, specifically F8-C4, are the regions most associated with MDD. Low PC scores for this connection are also associated with the MDD group, suggesting a disruption in the connectivity of brain networks that affect emotional regulation, cognitive processing and social cognition.

The second most important connection (Fp1-Fp2) associated with MDD is located in the frontopolar region of the brain, in both the right and left hemispheres. These regions are associated with affective processing, social cognition, working memory and perception. Low SC scores for this connection are associated with MDD patients and may be related to poor concentration, memory and decision-making difficulties. It may also contribute to problems in regulating emotions, leading to mood swings. These features are often observed in people with MDD.

Abnormal EEG patterns were found to be prevalent in both hemispheres of the MDD brain, which is consistent with existing literature. Interestingly, the application of complex network measures showed inferior classification performance compared to connectivity matrices. The test set yielded an AUC of 0.875 and an accuracy of 0.804. However, the network approach highlighted the importance of some measures such as assortativity, entropy of the degree distribution and density of connections. The functional MDD brain network has a more homogeneous degree distribution, resulting in a less structured topology, which could reduce the efficiency of information flow.

Our future study could include comparing local and global data to identify the most effective approach for patient differentiation and topology interpretation. In addition, exploring the different frequencies of EEG signals may provide insights into potential improvements in predictive accuracy and a deeper understanding of the intricate dynamics of MDD-related brain changes.

## Data Availability

All data produced are available online at: https://figshare.com/articles/dataset/EEG_Data_New/4244171

## VII. ACKNOWLEDGEMENTS

L.F.S. is indebted to Capes (grant 88887.645667/2021-00) for the financial provided to this research. F.A.R acknowledges CNPq (grant 308162/2023-4) and FAPESP (grants 24/02322-0, 20/09835-1 and 13/07375-0) for the financial support given for his research.

## Notes

### Competing Interest Statement

The authors have declared no competing interest.

### Author Declarations

The source data was openly available before the initiation of this study. It can be located in this link: https://figshare.com/articles/dataset/EEG_Data_New/4244171 For further details, see: https://www.sciencedirect.com/science/article/pii/S1746809416300866#sec0020

